# County Demographics and COVID-19 Death Rates: Comparison of relationship in the first and current stage of the pandemic in the United States of America

**DOI:** 10.1101/2020.11.24.20237818

**Authors:** Emma Urbine, Madison T James, Vishal Shah

## Abstract

Previous studies have discovered disparity in death rates associated with SARS-CoV-2 infection in the United States during the initial stages of the pandemic. Specifically, the death rates were higher in the population in poverty and communities of color across the United States. In the current study, we perform the secondary analysis of death rates due to COVID-19 data, obtained from the Center for Disease Control and Prevention (CDC). Results indicate that in the first phase of the pandemic (February 1 to August 1, 2020), counties with higher percentage of White, Native Hawaiian and other Pacific Islanders, and two or more races populations were found to have lower per-capita COVID-19 death rate. Whereas counties with population having higher percentage of females, Black or African American people, and persons in poverty had higher death rates. Analysis of the death rates from August 1 to September 10, 2020, indicate that disparity continues with counties having higher population of Black or African American people and female having higher death rates. Poverty is not a significant variable in determining the death rates due to COVID-19. Based on the current data and lack of detailed molecular mechanism of the disease, we suggest that more resources must be diverted to counties with higher percentages of Black or African American and female populations.

## Introduction

SARS-CoV-2 infections in the United States of America has taken a significant toll on the society with over six million positive cases and a death toll of over 180,000. However, the disease has not impacted the population uniformly. Studies on the disparity of the disease in the United States have shown that the disease had taken a larger toll in communities of color, and among under-resourced communities.^1,2^ Studies conducted within the first few months of the pandemic showed that Black and American Indian populations, as well as people living in low-income households suffer higher fatality rates due to COVID-19.^1,3,4^ The death toll in men was also higher than women due to COVID-19.^5^ However, no follow up studies have been conducted to investigate if the disparity in the death toll has continued in the population as the pandemic has progressed.

In the current study, using the county wide COVID-19 death toll data provided by the Center for Disease Control and Prevention (CDC) from February 1, 2020 to August 1, 2020 we investigate if the prior conclusions hold true. Then using the data of COVID-19 death toll from August 1, 2020 to September 16, 2020 we investigate if the conclusions still hold true.

## Methodology

### Data source

Data on Provisional COVID-19 deaths by County in USA were obtained from the CDC database.^6^ The population and demographic data for each county were obtained from the U.S. Census Bureau database.^7^ The demographic variables (independent variables) included were i) sex (percentage female population), ii) race (percentage of White, Black or African American, Asian, Native Hawaiian and other Pacific Islanders, American Indian and Alaska Native, Hispanic or Latino, and two or more races), and iii) income and poverty (median household income, per capita income, and percentage person in poverty).

The peak date of the second wave of COVID-19 pandemic in the United States was on July 19, 2020, with over 66,000 new cases of COVID-19 reported on that day. Adding two weeks to the date and looking at the data set from the CDC closest to the cut-off date, August 1, 2020 was selected as the date to separate what is referred in the study as the first phase of the pandemic and the second phase.

### Data transformation and analysis

The raw death numbers per county were transformed to death counts per 100,000 people and was the dependent variable. Transformation of data was performed by using the formula LN(x+1). Poisson regression analysis was performed using Minitab 18 software. Variables having *p* value <0.05 were considered to be statistically significant.

## Results and Discussion

Of the 3,141 counties and county equivalents in the United States, 799 of the counties had suffered from COVID-19 death toll between February 1 and August 1 of 2020. Majority of the counties having high per capita death toll were in the New York city metro region and along the I-95 corridor in Northeast (Table 1).

**Table 1.**
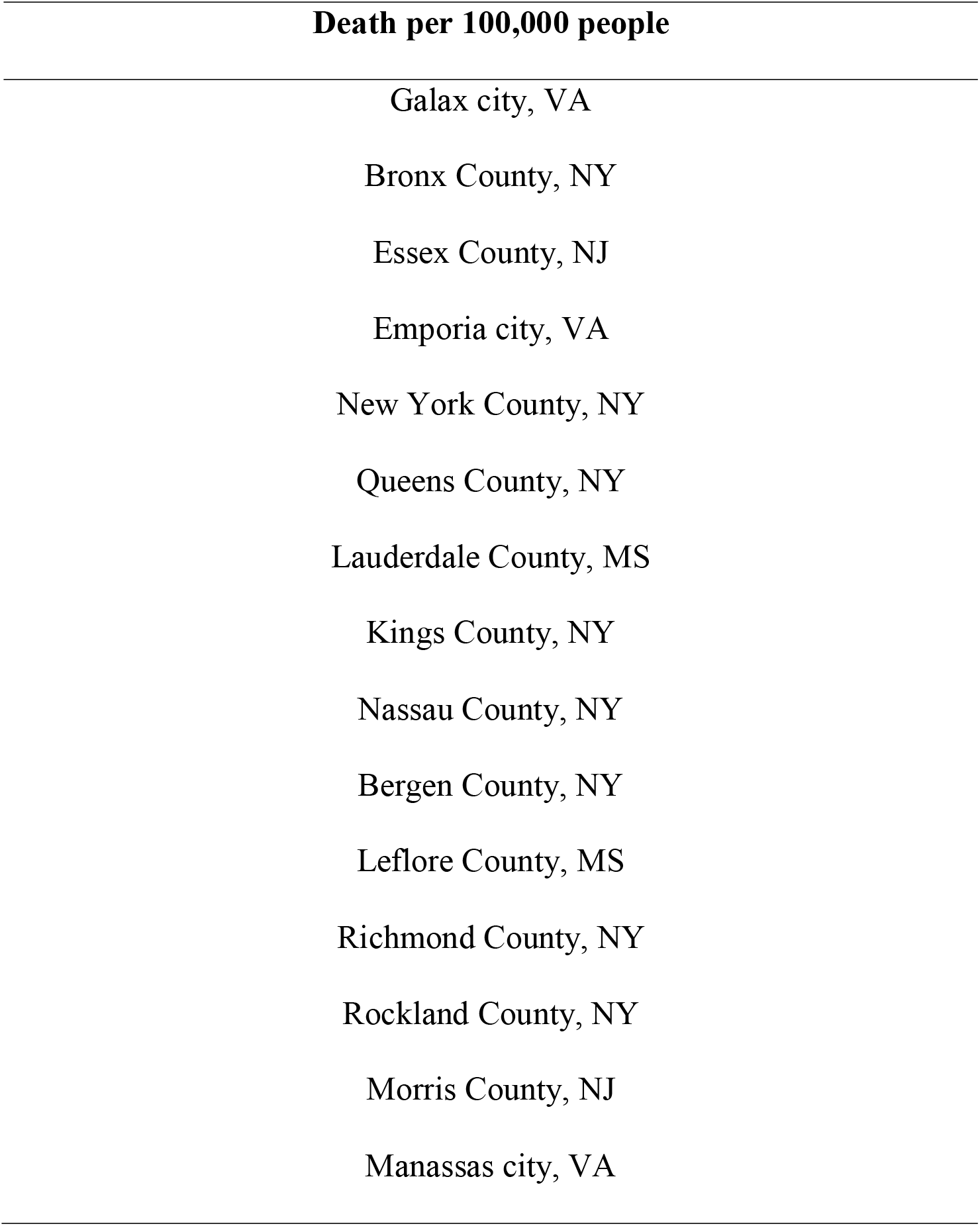
Top 15 counties in the United States showing highest death rate due to COVID-19 per 100,000 people from February 1, 2020 to August 1, 2020. The counties are sorted in descending order.

| Death per 100,000 people |
| --- |
| Galax city, VA |
| Bronx County, NY |
| Essex County, NJ |
| Emporia city, VA |
| New York County, NY |
| Queens County, NY |
| Lauderdale County, MS |
| Kings County, NY |
| Nassau County, NY |
| Bergen County, NY |
| Leflore County, MS |
| Richmond County, NY |
| Rockland County, NY |
| Morris County, NJ |
| Manassas city, VA |

Table 2 indicates the demographic variables that are statistically significant and have *p* values less than 0.05. Comparing the coefficients, counties having high White population, Native Hawaiian and other Pacific Islanders population, and population from two or more races had lower per-capita COVID-19 death rate. In contrast, counties having high female population, Black or African American population or people in poverty had higher per capita COVID-19 death rate. Demographic variables of percentage of Asian population (*p* = 0.43), Hispanic population (*p =* 0.56) and income and poverty variables of per capita income (*p* = 0.57) and median household income (*p* = 0.12) showed no statistical significance in the relationship to COVID-19 death rates.

**Table 2.**
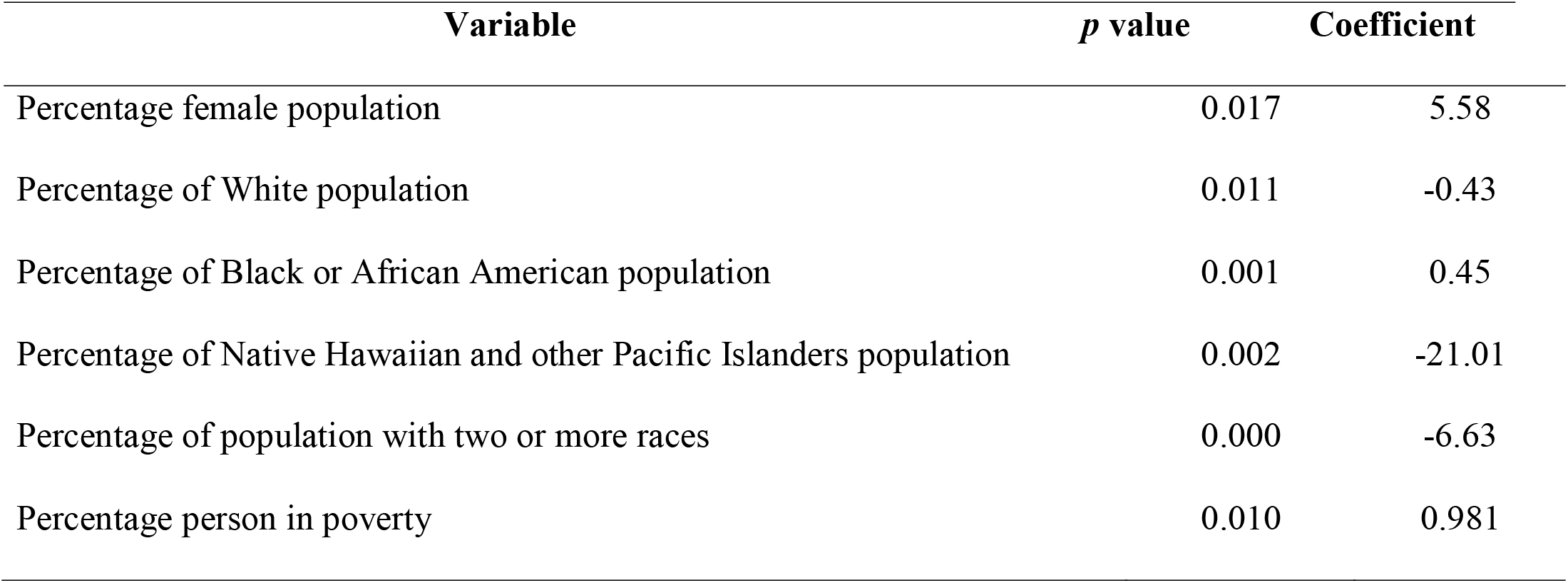
Variables showing *p* value < 0.05 in a Poisson regression model results for deaths from COVID-19 in the United States from February 1 to August 1, 2020.

| Variable | *p* value | Coefficient |
| --- | --- | --- |
| Percentage female population | 0.017 | 5.58 |
| Percentage of White population | 0.011 | -0.43 |
| Percentage of Black or African American population | 0.001 | 0.45 |
| Percentage of Native Hawaiian and other Pacific Islanders population | 0.002 | -21.01 |
| Percentage of population with two or more races | 0.000 | -6.63 |
| Percentage person in poverty | 0.010 | 0.981 |

The observed results are in line with those reported in the literature. During initial stages of pandemic, counties with higher poverty had large number of confirmed cases.^2^ Similarly, African American population have increased death rates due to COVID-19 then other populations. Various sociological and physiological parameters including availability of testing sites, co-morbidity, lack of trust in medical system, increased population as essential workers are some of the factors suggested to be behind the increased death toll in African American population.^8-11^

A positive correlation between increased per capita COVID-19 death rate and percentage of female population in a county has not been reported in the literature. The observation runs in contrast to the reported disparity of males being at greater risk of severe COVID-19 outcomes than women.^12^ Both sex (i.e., biological differences) and gender (i.e., sociocultural and behavioral differences) have been suggested to be playing a role in the observed disparity.^12^ At this stage, we are not in a position to propose the possible reasons behind the observed correlation of increased death rates in counties with higher female population. Further studies are urgently warranted to elucidate the factors governing the correlation.

Between August 1 and September 16, 2020, 213 additional counties reported death toll due to Covid-19. As can be seen in Table 3, majority of the top 15 counties reporting highest per capita death rate are in the states of Georgia, Virginia, and Texas.

**Table 3.** Top 15 counties in the United States showing highest death rate due to COVID-19 per 100,000 people from August 1, 2020 to September 16, 2020. The counties are sorted in descending order.

| Death per 100,000 people |
| --- |
| Seminole County, FL |
| Richmond city, VA |
| Martinsville city, VA |
| Crenshaw County, AL |
| Little River County, AR |
| Newton County, AR |
| Pulaski County, GA |
| Jenkins County, GA |
| Toombs County, GA |
| Somervell County, TX |
| Cameron County, TX |
| Laurens County, GA |
| Candler County, GA |
| Hidalgo County, TX |
| Charlottesville city, VA |

When one performs Poisson regression analysis between the per capita death rates for this period and the variables described in Table 1, only four variables out of the six were found to be statistically significant (Table 4). Percentage of Native Hawaiian and other Pacific Islanders (p = 0.40) and the percentage of population in poverty within a county (*p* = 0.18) were not found to be statistically significant during this analysis. This indicates that while COVID-19 death rate may have been higher during the initial stages of the pandemic in the population living in poverty, this is not the case in the recent stage of the pandemic. Initial lag in testing and availability of health care to the population living in poverty may have been corrected over time, resulting in death rates similar to the general population.

**Table 4.** Variables showing *p* value < 0.05 in a Poisson regression model results for deaths from COVID-19 in the United States from August 1 to September 16, 2020.

| Variable | *p* value | Coefficient |
| --- | --- | --- |
| Percentage female population | 0.001 | 6.74 |
| Percentage of White population | 0.000 | -0.67 |
| Percentage of Black or African American population | 0.001 | 0.47 |
| Percentage of population with two or more races | 0.004 | 4.22 |

Of concern, is the continuing trend of the disparity in the per-capita death rates amongst Black and African American population seems to be a signature of the pandemic and needs urgent attention. In contrast, throughout the pandemic thus far, counties having higher White population have lower per capita death rates. Earlier studies have shown that individuals in predominantly non-white communities were nearly 8 times more likely to become infected and 9 times more likely to die due to COVID-19 infections.^13^ Historically, disparities in health outcomes are not new in the United States but are rooted in structural inequities. COVID-19 pandemic has resulted in these disparities precipitating out to our full glare. Millet et al. have recently suggested that individual level-characteristics such as presence of co-morbidity do not fully explain excess disease burden in racial minority communities. The problem and eventual solution lies at the structural and policy-level.^14^

Thus far, the approach for combatting the pandemic across the United States has been one size fits all. The underlining approach has been to mitigate the spread across the entire country. While the approach has its merits, when resources such as testing for SARS-CoV-2 and contact tracing are limited, it is imperative to increase the resources for the populations that would be more susceptible to prevent increased casualty across the country. This is more critical since we do not understand many aspects of the molecular mechanism of the disease and the underlining biology behind the disparity. Based on the observed results, we call upon the policy makers to divert more resources to counties having higher population of Black and African Americans and those having higher percentage of female citizens. The resources should not just be limited to testing, contact tracing, and availability to medical care. With increased death toll in these populations, there are many negative impacts on the emotional well-being of the citizens. The overall community grief due to the loss of loved ones is compounded by the lack of community grieving process due to the restrictions on gathering due to the pandemic.^15^ This grief compounds the emotional pain and further increases the physical illness burden. Over long term this could lead to further economic hardship that exacerbates the disparity. Community based organizations, employers, public health agencies, and policy makers need to increase the availability of resources to manage the mental health care of the population suffering excess grief.

## Data Availability

See references.

